# Classifications of Breast Cancer Images by Deep Learning

**DOI:** 10.1101/2020.06.13.20130633

**Authors:** Wenzhong Liu, Hualan Li, Caijian Hua, Liangjun Zhao

## Abstract

**Background:** Breast cancer is a leading cause of cancer-related death in women. Classifications of pathological images are important for its diagnosis and prognosis. However, the existing computational methods can sometimes hardly meet the accuracy requirement of clinical applications, due to uneven color distribution and subtle difference in features.

**Methods:** In this study, a novel classification method DeepBC was proposed for classifying the pathological images of breast cancer, based on the deep convolution neural networks. DeepBC integrated Inception, ResNet, and AlexNet, extracted features from images, and classified images of benign and malignant tissues.

**Results:** Additionally, complex tests were performed on the existing benchmark dataset to evaluate the performance of DeepBC. The evaluation results showed that, DeepBC achieved 92% and 96.43% accuracy rates in classifying patients and images, respectively, with the F1-score of 97.38%, which better than the state-of-the-art methods.

**Conclusions:** These findings indicated that, the model had favorable robustness and generalization, and was advantageous in the clinical classifications of breast cancer.

## Introduction

Breast cancer is one of the most common malignant diseases that affect female health, which is linked with high morbidity and mortality [11]. The optimal treatment for breast cancer depends on sophisticated classification. Early diagnosis and classification of breast cancer are conducive to the option of therapeutic methods and the control over tumor cell metastasis, and this has significantly improved the patient survival rate [40]. Doctors who understand the breast cancer types can develop targeted treatment plans based on the unusual clinical manifestations and prognostic outcomes of various breast cancers. In short, classification treatment of breast cancer lays the foundation for accurate medical planning, which is of great significance for clinical diagnosis and prognosis[8].

Breast cancer can be diagnosed through breast ultrasound and breast biopsy, among which, biopsy is the only diagnostic approach to determine whether a suspicious area is cancerous [6]. For each patient, the breast tissue-derived cells are collected onto a glass slide for hematoxylin and eosin (H&E) staining by the pathologist [23]. Microscopic analyses of breast tissue slides at different magnifications are carried out to identify two types of lesions, namely, benign and malignant [2]. Benign tumors are comprised of abnormal epithelial cells and are not cancerous, and most epithelial cells will not develop into breast cancer. Malignant tumors or cancer cells are cells that begin to divide abnormally and grow without rules [25]. Analyzing microscopic images is a complex, time-consuming and difficult task, and the excessive fatigue can result in misdiagnosis [14]. Therefore, only the trained pathologists are responsible for analyzing the histopathological images[1]. However, skilled pathologists are lacking in underdeveloped areas and small hospitals, such as primary hospitals and clinics [37]. Therefore, it is important to develop an automated system to distinguish the cancerous tissue (malignant tissue) from the non-cancerous tissue (benign tissue) [33]. Such a system should help pathologists to simplify the diagnosis, save time, and increase the diagnostic efficiency.

In most automated systems, traditional machine learning-based methods are adopted to extract the texture, morphology, and structural features of cell image or cell nucleus[17]. Afterwards, the features built on images and textures are obtained and utilized to classify different pathological images and to establish an automated system. Some researchers analyze the nucleus by extracting the nucleus characteristics [21], thus providing essential information for classifying benign and malignant cells[39]. Notably, the methods for manual feature extraction include thresholds, clustering, active contours, watersheds, and graphic cutting. Based on the traditional machine learning algorithms, different nucleus segmentation algorithms are employed for feature descriptors, such as support vector machines and random forests [35]. Using these algorithms, the distinguishing features are extracted from the nucleus to distinguish the benign slides from the malignant one. An improved morphological feature to discover the single-cell regions using wavelet transforms is also applied in nucleus classification. Typically, nucleus segmentation and classification using clustering algorithms and Hough transform [12, 13] represent the similar principles. The active contour technology segments cells from the background of breast histopathological images. This method extracts the nucleus morphological features and divides them into two types, which are normal cells and tumor cells[15]. The watershed segmentation technology extracts nucleus information from breast cancer histopathological images. Noteworthily, most classification methods are performed on low-resolution images with different magnifications.

The deep learning models are employed to solve the classification problems in breast cancer detection[34]. Deep learning is a non-linear representation learning method, which belongs to machine learning. Convolution neural network (CNN), a kind of deep learning, becomes a general-purpose feature extractor. CNN classifies the histopathological images of breast cancer with independent magnification, thus obtaining a higher recognition rate[10, 24]. In addition, CNN can more accurately detect breast cancer metastasis, which helps the pathologists to make a diagnosis[3]. For example, AlexNet divides the pathological images of breast cancer into two categories: benign and malignant [28], and achieves higher accuracy rate than those of traditional machine learning algorithms. Besides, ResNet and Inception are the fast and accurate models for cell-based image classification [16]. Among the various machine learning algorithms with artificial features, the most attractive feature about CNN is that there is no need to design the feature extraction methods [22]. However, the number of features generated by CNN may be significant, which may result in overfitting and information redundancy [7]. Consequently, the accuracy of the system gradually decreases with the increase in magnification.

To accurately and reliably solve the classification problem of breast cancer, a deep CNN, namely, the DeepBC model, was proposed. DeepBC was able to classify two types of pathological images of breast cancer. This article focused on classifying the malignant and benign tumor biopsy images using the BreaKHis dataset. Moreover, the DeepBC model was rigorously evaluated. Our evaluation results showed that, DeepBC achieved the accuracy rates of 92% and 96.43%, respectively, in distinguishing patients and images, with the F1-score of 97.38%, and DeepBC attained superior efficiency to other methods.

## Methods

### BreakHis dataset

In this study, BreakHis [29], the breast cancer dataset of microscopic images, was utilized to evaluate the performance of DeepBC. BreakHis contains 7,909 breast cancer biopsy images at different microscopic magnifications (×40, ×100, ×200, and ×400). Each pathological image is a 700×460 pixel png format file with 3 RGB channels. Later, BreakHis was stained with hematoxylin and eosin (H&E), and images were divided into two categories: malignant and benign. A total of 2480 samples were included, including four types of benign breast tumors (namely, adenosis A, fibroadenoma F, phyllodes tumor PT, and tubular adenona TA), and four types of malignant tumors (breast cancer: carcinoma DC, lobular carcinoma LC, mucinous carcinoma MC, and papillary carcinoma PC). In this study, 70% images were employed for training, while the remaining 30% were utilized for independent testing. Each type of image was allocated with a training set and a test image ratio of 7: 3.

### Input layer

This layer loads breast cancer images from the database and outputs to fill the convolutional layer. The input image is a pathological tissue image with a size of 227 × 227, which consists of three 2-D arrays with the red, green, and blue channels.

### Convolutional layer

The convolutional layer performs the convolution operation on input data through a filter to create a feature map. A filter represents the weight of a two-dimensional matrix shared by neurons, and the common filter kernels are 1×1, 3×3, 5×5, 7×7, 9×9, and 11×11. Then, the distance between two neighboring patches is called stride. A filter slides along the height and width of the input image, calculates the dot product, and produces a two-dimensional feature map. Afterwards, the convolutional layer sends all local weight values to the next layer by rectified linear activation (ReLU), a non-linear function to avoid linearization of the convolutional layer [19]. The gradient of activation function is larger, whereas the optimization of loss function is simpler and more effective.

### Pooling layer

The pooling layer mainly functions to combine the semantically similar features into one feature, and its output remains the same ratio, but the number of parameters decreases. The pooling layer reduces the size and noise, which thereby accelerates the convergence speed and improves the model generalization performance. There are three strategies in the pool layer, namely, maximum pooling, average pooling, and random pooling. The MaxPool layer [9] is used in this architecture, and the maximum value of patches is calculated by merging units.

### Inception module

The Google network model holds the Inception architecture for large-scale visual recognition challenges [32]. Inception architecture is a method that gradually increases the feature size, and saves the depth of CNN without increasing the computational memory. Each Inception architecture can normalize data, and decomposes a large convolutional layer into a series of parallel convolutional layers [31]. The convolution kernel is limited to 1 ×1, 3 ×3, and 5 ×5.

### ResNet module

The generally stacked CNNs tend to overfit the training data and perform poorly on the actual data. In our model, the ResNet layer was employed to address the degradation problem of deep learning networks. The ResNet network turns several layers in the original network into a residual block[38], which accelerates the training speed of deep networks, and renders faster network convergence.

### AlexNet module

The AlexNet [18] module contains 5 convolutional layers and 2 pooling layers. There are 96 (11 ×11), 256 (5 ×5), and 384 (3 ×3) filters in the first, second and the last three convolution layers, respectively. Meanwhile, there is a ReLU activation function after each convolutional layer, and the maximum pooling layer follows the first and second convolutional layers.

### Fully connected layer

Although the convolutional layer can obtain multi-scale information from the input data, the connection between the feature map and the classification score remains a black box. The fully connected layer declares many hidden layers with specified neurons and activation functions, and these hidden layers map the convolution features into a classification score.

When the number of feature maps or trainable parameters is equal or greater than that of samples, there is a risk of overfitting. To avoid overfitting, the number of features should be much lower than that of samples. The dropout layer [30] is located between two fully connected layers, and some neurons are discarded with the probability value of p=0.5. The dropout layer can reduce excessive merging behavior, so as to improve the CNN classifier performance.

### Structure of the DeepBC model

The DeepBC deep learning model helped to better understand the inherent laws and characteristics of breast cancer. In this study, the network structures of Inception, ResNet and AlexNet were adopted. In the fields of medicine and digital pathology, AlexNet is representative of CNN. Inception structure is a parallel convolutional layer that contains singular convolution kernels, while ResNet is an advanced network structure to calculate residuals block. Figure 1 shows the DeepBC network architecture. The architecture of DeepBC was constructed through a series of stages, including the input layer, the “Inception + ResNet + AlexNet” structures. The fully connected layer had 512 neuron nodes and two output classes. The DeepBC realized the transfer of features from low-level to more abstract high-level, which was beneficial to learn the feature rules inherent in the pathological image.

**Figure 1.**
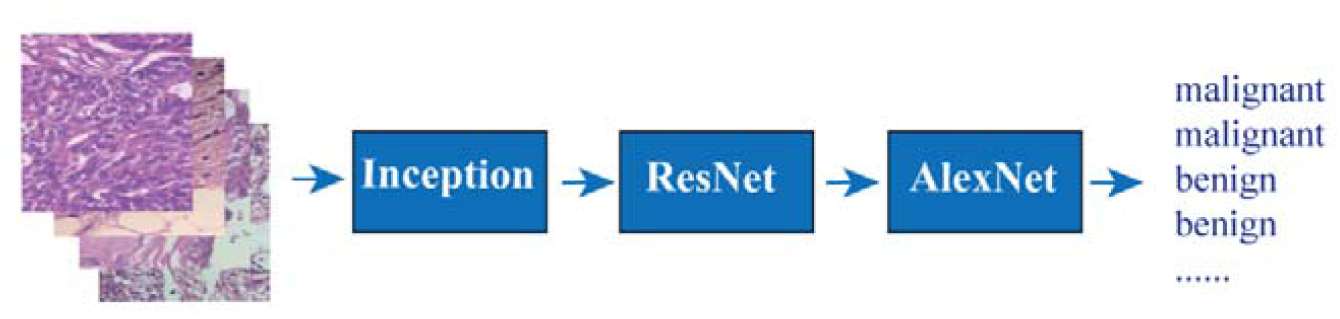
the architecture of DeepBC.

As observed in Figure 1, DeepBC directly trained or evaluated the BreakHis dataset. By learning the hierarchical features in the training model, DeepBC fitted the parameters of weight and bias. Then, DeepBC started learning the underlying features, such as color, texture, and shape, after inputting the breast cancer pathological images to the input layer. Later, DeepBC extracted the semantic features and sent them to the fully connected layers for classification through progressively learning “Inception +ResNet + AlexNet”.

### Training strategy

In this study, no preprocessing, dimensionality reduction or data enhancement method was employed before the image was input into the network. This experiment was conducted by Nvidia GTX1060 GPU equipped with 6GB RAM. The Pytorch package was the deep learning framework for implementing DeepBC. The size of each input image was resized to 227 ×227 pixels, and the epoch was 78, with Adam optimizer and 10e-4 learning rate. We used each epoch-trained model to predict the test set respectively, and selected the prediction probability file with the highest accuracy. It took nearly one day to train and predict.

### Evaluation metrics

To quantify and compare our results, the following evaluation metrics were adopted:

1. **Patient accuracy**. Mean patient level accuracy [5] is defined as:

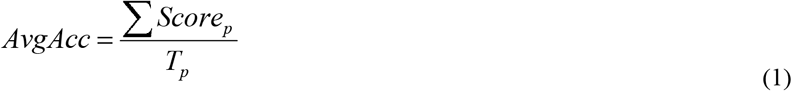 Where Tp stands for the total number of patients in the test set. Patient score is defined as follows:

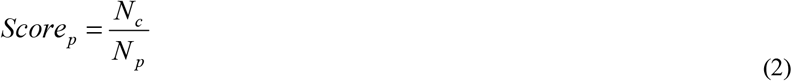 Where Np represents the number of biopsy images of patient P, and Nc indicates the correctly classified frames of images for patient P.
2. **Image accuracy** [5]. Image accuracy is calculated at the image level (in other words, patient information is not taken into consideration), which represents a method to evaluate the accuracy of CNN model image classification. Image accuracy is defined as follows,

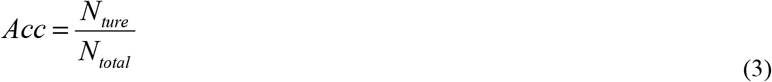 Where *N*_*true*_ is the number of correct classifications, and *N*_*total*_ represents the total number of pictures.
3. **F1-score** [27]. F1-score is adopted to better highlight the sensitivity to (positive) malignancy, which is defined as the harmonic mean between sensitivity (also called Recall) and Precision [4]:

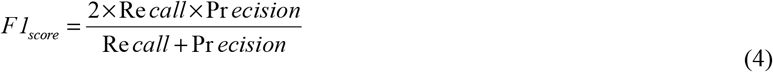 Where

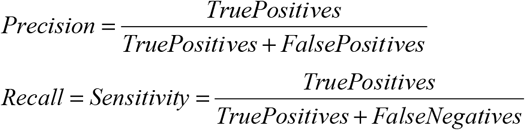
4. **Misclassification rate**. The ratio is the number of misclassified images in each sub-category to the total number of each sub-category.

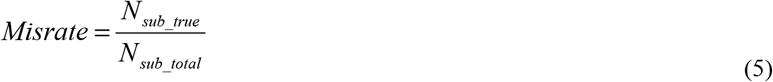 Where *N*_*sub*_*true*_ is the number of misclassified images in each sub-category, *N*_*sub*_*total*_ is the total number of each sub-category.

## Results

### Confusion matrix and misclassification rate

To evaluate the model performance, the confusion matrix was utilized in this study to describe the classification results of the test set. As shown in the confusion matrix (Figure **2**), the benign and malignant tumors achieved the high classification accuracy rates of 95.83% and 96.71%, respectively. However, 4.17% malignant tumors were incorrectly classified as benign, whereas 3.29% benign tumors were wrongly classified as malignant.

**Figure 2.**
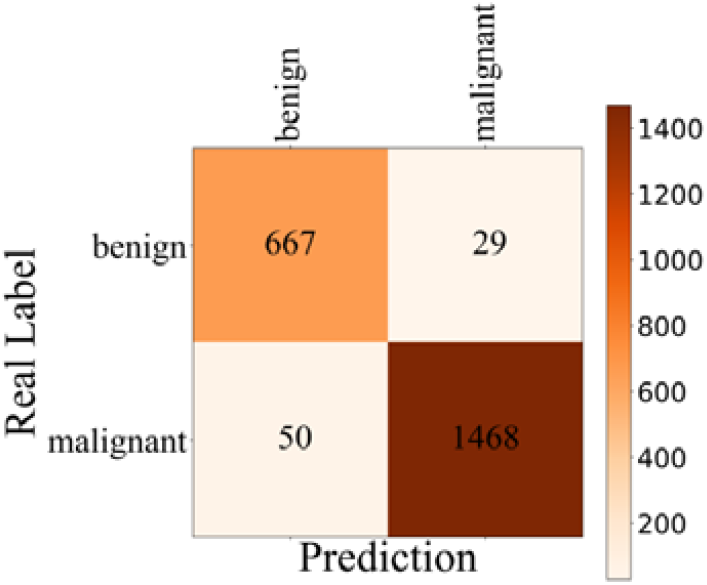
confusion matrix.

We compared the subclass misclassification rates of benign and malignant (Table 1). Table 1 shows misclassification rates of the benign and malignant for DeepBC were meager compared with other tools, but VGG16 and AlexNet were higher. It was worth noting that the misclassification rate of fibroadenoma and lobular carcinoma by the GoogLeNet was slightly lower than the DeepBC. For all methods, the misclassification rate of benign was generally higher than the malignant. It indicated that the relatively low missed diagnosis rate for malignant tumors, the misdiagnosis rate for benign tumors was relatively high. The reason might be ascribed to the few clinically records of benign subtypes. Therefore, the DeepBC model attained a higher sensitivity and performed better in the detection and classification of breast cancer.

**Table 1.** misclassification rates of several methods

|  | Tumor Type | DeepBC | GoogLeNet | ResNet | VGG16 | AlexNet |
| --- | --- | --- | --- | --- | --- | --- |
| benign | adenosis | 0.79 | 10.32 | 10.32 | 15.87 | 20.63 |
|  | fibroadenoma | 3.17 | 2.82 | 7.39 | 22.54 | 21.13 |
|  | phyllodes tumor | 7.75 | 10.85 | 16.28 | 33.33 | 31.78 |
|  | tubular adenoma | 5.73 | 12.10 | 14.65 | 30.57 | 28.03 |
| malignant | ductal carcinoma | 1.04 | 1.35 | 4.36 | 2.49 | 3.22 |
|  | lobular carcinoma | 5.00 | 3.33 | 10.00 | 6.11 | 4.44 |
|  | mucinous carcinoma | 10.96 | 10.50 | 9.59 | 24.20 | 21.46 |
|  | papillary carcinoma | 4.52 | 8.39 | 10.97 | 21.94 | 16.13 |

### Classification comparison of accuracy and F1-score

We used the sklearn.metrics package to calculate the accuracy and F1-score. The overall accuracy rates of DeepBC for images and patients were 96.43% and 92.00%, respectively, with the F1-score of 97.38%, which was higher than those obtained by other tools (Table 2). We further focused on the impact of magnification on the classification accuracy. Firstly, at the magnification of 40 ×, the accuracy rates for images and patients achieved by DeepBC were 97.31% and 100.00%, respectively with the F1-score of 98.03%, which was superior to those by other tools(Table 3). When the magnification was 100x, the accuracy rates of images and patients attained by DeepBC were 96.06% and 83.33%, respectively, and the F1-score was 97.11% (Table 4), which was superior to those obtained by other tools. At the magnification of 200x, the accuracy rates of images and patients achieved by DeepBC were 97.53% and 100.00%, respectively, and the F1-score was 98.21%(Table 5), which outperformed other methods. When the magnification was 400x, the accuracy rates of images and patients achieved by DeepBC were 94.66 % and 85.71%, respectively, and the F1-score was 96%(Table 6). According to the above results, the feature learning ability of DeepBC was much better than the state-of-the-art classifiers.

**Table 2.** overall accuracy rates of several methods

| Method | Accuracy<br>(Image) | Accuracy<br>(Patient) | F1-score |
| --- | --- | --- | --- |
| DeepBC | 96.43 | 92.00 | 97.38 |
| GoogLeNet | 95.08 | 92.00 | 96.41 |
| ResNet | 92.05 | 72.00 | 94.16 |
| VGG16 | 86.59 | 56.00 | 90.39 |
| AlexNet | 87.26 | 84.00 | 90.89 |

**Table 3.**
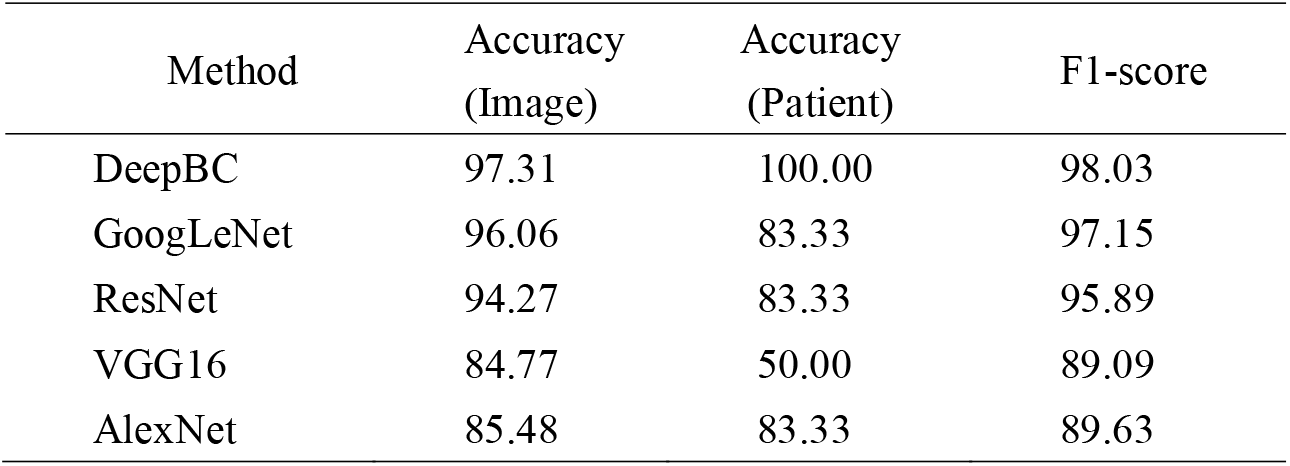
accuracy rates of several methods at the magnification of 40 ×

| Method | Accuracy<br>(Image) | Accuracy<br>(Patient) | F1-score |
| --- | --- | --- | --- |
| DeepBC | 97.31 | 100.00 | 98.03 |
| GoogLeNet | 96.06 | 83.33 | 97.15 |
| ResNet | 94.27 | 83.33 | 95.89 |
| VGG16 | 84.77 | 50.00 | 89.09 |
| AlexNet | 85.48 | 83.33 | 89.63 |

**Table 4.** accuracy rates of several methods at the magnification of 100 ×

| Method | Accuracy<br>(Image) | Accuracy<br>(Patient) | F1-score |
| --- | --- | --- | --- |
| DeepBC | 96.06 | 83.33 | 97.11 |
| GoogLeNet | 95.38 | 83.33 | 96.60 |
| ResNet | 93.15 | 83.33 | 94.96 |
| VGG16 | 86.82 | 83.33 | 90.62 |
| AlexNet | 86.64 | 83.33 | 90.39 |

**Table 5.** accuracy rates of several methods at the magnification of 200 ×

| Method | Accuracy<br>(Image) | Accuracy<br>(Patient) | F1-score |
| --- | --- | --- | --- |
| DeepBC | 97.53 | 100.00 | 98.21 |
| GoogLeNet | 95.41 | 100.00 | 96.72 |
| ResNet | 92.05 | 66.67 | 94.25 |
| VGG16 | 87.81 | 33.33 | 91.43 |
| AlexNet | 87.63 | 83.33 | 91.34 |

**Table 6.** accuracy rates of several methods at the magnification of 400 ×

| Method | Accuracy<br>(Image) | Accuracy<br>(Patient) | F1-score |
| --- | --- | --- | --- |
| DeepBC | 94.66 | 85.71 | 96.00 |
| GoogLeNet | 93.28 | 100.00 | 94.99 |
| ResNet | 88.34 | 57.14 | 91.07 |
| VGG16 | 86.96 | 57.14 | 90.35 |
| AlexNet | 89.53 | 85.71 | 92.37 |

## Discussion

The computer-assisted biopsy analysis is mainly carried out to minimize the manual operation of microscopically stained slides [26]. Using the image processing and machine learning technology, a classification computer-aided system can automatically diagnose breast cancer. In recent years, breakthroughs have been made in primary medical image analysis and detection methods. Many researchers have established the computer-aided systems to automatically classify benign and malignant tissue images. This improves the diagnosis efficiency, reduces the tedious work of pathologists, and avoids the possibility of misdiagnosis. Fortunately, the mortality of breast cancer dramatically reduces, even though its incidence significantly increases.

However, the color appearance changes in H&E stained histopathological images have afflicted researchers in automatic image analysis. The benign and malignant cells show irregular morphology, excessive overlapping, and uneven color distribution [36]. The histopathological images of breast cancer are unique in different classifications, but the pathological images of the same tumor tissues also have significant differences in features such as resolution, contrast, and appearance, making it difficult to distinguish breast cancer. Notably, pathologists have quickly adapted to various color changes without affecting the diagnosis accuracy. However, the color variability has adversely affected computer analysis[20]. Many approaches can be utilized to standardize colors, among which, color standardization becomes an essential tool for the quantitative analysis of histopathologically stained color images. However, these approaches depend on artificial methods to extract features, which needs great efforts and expertise and thus severely restricts the application of machine learning methods in the histopathological classification of breast cancer. In this study, the DeepBC model does not require color standardization of the image, and it also extracts features of the pathological tissue image. Typically, our proposed method extracts not only the low-level features but also the representative features, which is suitable for classifiers with discriminatory analysis abilities. Based on findings in this study, DeepBC displays useful application prospects.

## Conclusion

Breast cancer is a leading cause of cancer-related death in women. Multiple classifications are important for its diagnosis and prognosis. In this study, to accurately and reliably solve the multi-classification problem of breast cancer, a deep CNN, namely, the DeepBC model, was proposed. DeepBC integrated AlexNet, ResNet, and Inception, extracted features from images, and classified images of benign and malignant tissues. DeepBC was able to classify multiple types of pathological images of breast cancer. This article focused on classifying the malignant and benign tumor biopsy images using the BreaKHis dataset. Moreover, the DeepBC model was rigorously evaluated. Our evaluation results showed that, DeepBC achieved the accuracy rates of 92% and 96.43%, respectively, in distinguishing patients and images, with the F1-score of 97.38%, and DeepBC attained superior efficiency to other methods. These findings indicated that, the model had favorable robustness and generalization, and was advantageous in the clinical multiple classifications of breast cancer.

## Data Availability

The datasets supporting the conclusions of this article are available at https://github.com/lwzyb/DeepBC.

## Compliance with Ethical Standards

### Consent for publication

Not applicable.

### Funding

This work was funded by a grant from the Natural Science Foundation for Talent Introduction Project of Sichuan University of Science and Engineering (award number 2018RCL20, grant recipient WZL).

### Conflict of Interest

The Author wenzhong Liu has received the research grant from Sichuan University of Science and Engineering. Huanlan Li declares that she has no conflict of interest. Caijian Hua declares that he has no conflict of interest. Liangjun Zhao declares that he has no conflict of interest.

### Ethical approval

This article does not contain any studies with human participants or animals performed by any of the authors. The BreakHis database used in this project is a public database. We have cited BreakHis’ work as required by the owner.

### Author’s contribution

Funding was obtained by WZL. WZL and HLL conceived the study. WZL designed the algorithms and the software, make performance evaluation, and wrote the manuscript. ZLJ and HCJ tested the software. The final version of the manuscript is approved by all authors.

## Acknowledgements

Not applicable.

## Author details

^1^ School of Computer Science and Engineering, Sichuan University of Science & Engineering, Zigong, 643002, China. ^2^ School of Life Science and Food Engineering, Yibin University, Yibin, 644000, China.

## Notes

### Competing Interest Statement

The authors have declared no competing interest.

### Author Declarations

The BreakHis database used in this project is a public database. We have cited BreakHis' work as required by the owner.

